# Discovery of Dynamic Models for AML Disease Progression from Longitudinal Multi-Modal Clinical Data Using Explainable Machine Learning

**DOI:** 10.1101/2025.04.07.25325267

**Authors:** Reza Mousavi, Moaath K. Mustafa Ali, Daniel Lobo

## Abstract

Acute Myeloid Leukemia (AML) is a complex and heterogeneous disease identified by severe clinical progression, fast cellular proliferation, and often high mortality rates. Incorporating diverse longitudinal information on patients’ medical histories is essential for developing effective disease predictive models applicable to both research and clinical settings. Here, we present a robust methodology for discovering dynamic predictive models to elucidate AML disease progression dynamics from a novel longitudinal multimodal clinical dataset of patients diagnosed with AML. The clinical dataset was analyzed to reveal the main clinical, genetic, and treatment features modulating disease progression. To discover mathematical models—including interactions, parameters, and nodes—predictive of AML progression, we present an explainable machine learning algorithm based on high-performance evolutionary computation. The results demonstrate that the predictive methodology could accurately estimate the clinical dynamics of AML progression in terms of blast percentages for both training and novel patients. This study demonstrates that the developed explainable machine learning approach can successfully predict AML progression by leveraging the heterogeneous and longitudinal dynamics of patients’ clinical data. More importantly, this methodology shows significant potential for application in modeling the progression dynamics of other acute diseases, providing a flexible and adaptable framework for advancing clinical and translational research.

## 1. Introduction

Acute myeloid leukemia (AML) is an aggressive and heterogeneous malignancy of the myeloid lineage, characterized by clonal expansion of abnormal precursor cells and impairment of normal hematopoiesis (1). AML is driven by different genetic and cytogenetic abnormalities within myeloid precursors that result in neoplastic transformation (2–4). Poorly differentiated myeloid cells can accumulate in the bone marrow, their site of origin, and circulate in the peripheral blood, causing complications such as an increased risk of infections, bleeding, fatigue, and bone pain (5, 6). The incidence of AML has continued to increase over the past few decades (7). The American Cancer Society estimates approximately 59,610 new cases and 23,710 deaths due to leukemia (all types) in the United States in 2023. Among these, approximately 20,380 new cases are expected to be diagnosed with AML, mostly in adults, resulting in 11,310 deaths. AML accounts for approximately 1% of all cancers and is more common in adults over the age of 45 years; however, AML can also occur in children. AML progresses rapidly, with a 5-year survival rate of approximately 28% in adults (8). Hence, the rapid and precise prediction of disease progression is essential for cost-effective treatment decisions and improving patient outcomes.

Owing to its significant contribution to predictive medicine and modern oncology, machine learning (ML) has recently gained increasing attention in cancer research (9–14). Recent studies have demonstrated that ML has excellent capabilities for handling large amounts of complex data and may prove to be a powerful tool for understanding and treating diseases (15–18). In AML, a highly heterogeneous hematologic malignancy, interpreting diagnostic tests, such as genetic mutations, chromosomal abnormalities, and blast percentages, has historically required experienced clinicians with years of training and expertise. However, the landscape of AML diagnosis and management has significantly changed with the advent of ML algorithms (19–21). Recent ML algorithms have been applied to several tasks in AML, including the initial diagnosis (22–24), marker and therapeutic target discovery (25, 26), and prognosis and treatment response estimation (27–31). However, existing ML models lack explainability, making it difficult for clinicians to understand how specific markers or interactions contribute to predicting disease prognosis. This limits the applicability of such models in clinical practice, where interpretability is critical for decision making. Furthermore, these ML approaches rely on static data and lack the capability to incorporate temporal information from patient medical records to predict disease progression. Such static models overlook the dynamic nature of AML, in which disease progression and treatment response evolve over time.

Dynamic mathematical models have been proposed to predict AML disease progression from clinical data (32, 33). These approaches can simulate the dynamics of leukemic cells during the clinical history of patients and predict disease prognosis (34, 35), treatment scheduling (36, 37), transplant dosage (38), and specific therapy response (39). However, several significant challenges remain. One major challenge lies in the complexity of managing and integrating high-dimensional, multimodal longitudinal clinical data into a mathematical dynamic model, which includes patient information, leukemia-associated genetic factors, and treatment responses collected at different time points. Conventional temporal dynamic methods struggle to effectively integrate and analyze these large and diverse data types, often leading to suboptimal results with limited clinical relevance. To address these gaps, innovative methodologies are needed that can efficiently process and analyze multimodal data on a large scale while providing explainable insights with dynamic models of AML progression.

Here, we propose an explainable machine learning methodology to derive complex dynamic models of AML progression from longitudinal multimodal data. This innovative method integrates the strengths of evolutionary computation and high-performance computing (40–44) to predict disease progression dynamics. The proposed methodology infers models that include both the topology and parameters of a dynamic system, represented by a system of ordinary differential equations (ODEs). By simulating these ODE-based models, the approach effectively recapitulates and predicts the observed disease progression dynamics from clinical data, thereby providing a framework for understanding and predicting AML trajectories. To evaluate the approach, dynamic models were inferred from a highly annotated, real-world AML clinical dataset collected at the University of Maryland Medical Center. These findings demonstrate the effectiveness of the methodology in capturing the longitudinal dynamics of AML progression and accurately predicting outcomes in new patients. This study establishes a foundation for advancing AML prognosis, personalized treatment strategies, and improving patient outcomes, with potential applicability in modeling the progression dynamics of other acute diseases.

## 2. Results

Leveraging large-scale clinical multimodal data alongside longitudinal aspects of patients’ medical records is essential for advancing personalized medicine to address unmet clinical demands in acute diseases. This approach facilitates a deeper understanding of disease progression and paves the way for the development of tailored interventions to improve patient outcomes. To this end, we present a computational framework that processes longitudinal multimodal data to infer dynamic models capable of predicting AML disease progression dynamics (Fig. 1). These interpretable models consist of a system of dynamic mathematical equations including quantitative input nodes, which include patient information, leukemia parameters, and treatment interventions, together with inferred intermediate nodes, and an output node, which predicts the temporal dynamics of disease progression—in the case of AML measured as the percentage of blasts abnormally present in the bone marrow or blood. The models also feature regulatory interactions, allowing nodes to positively and negatively affect other nodes. The intermediate and output nodes are capable of self-regulation and can regulate other nodes. However, the input nodes cannot be affected by regulations from other nodes. This design allows the models to capture the complex regulatory dynamics underlying disease progression.

**Figure 1.**
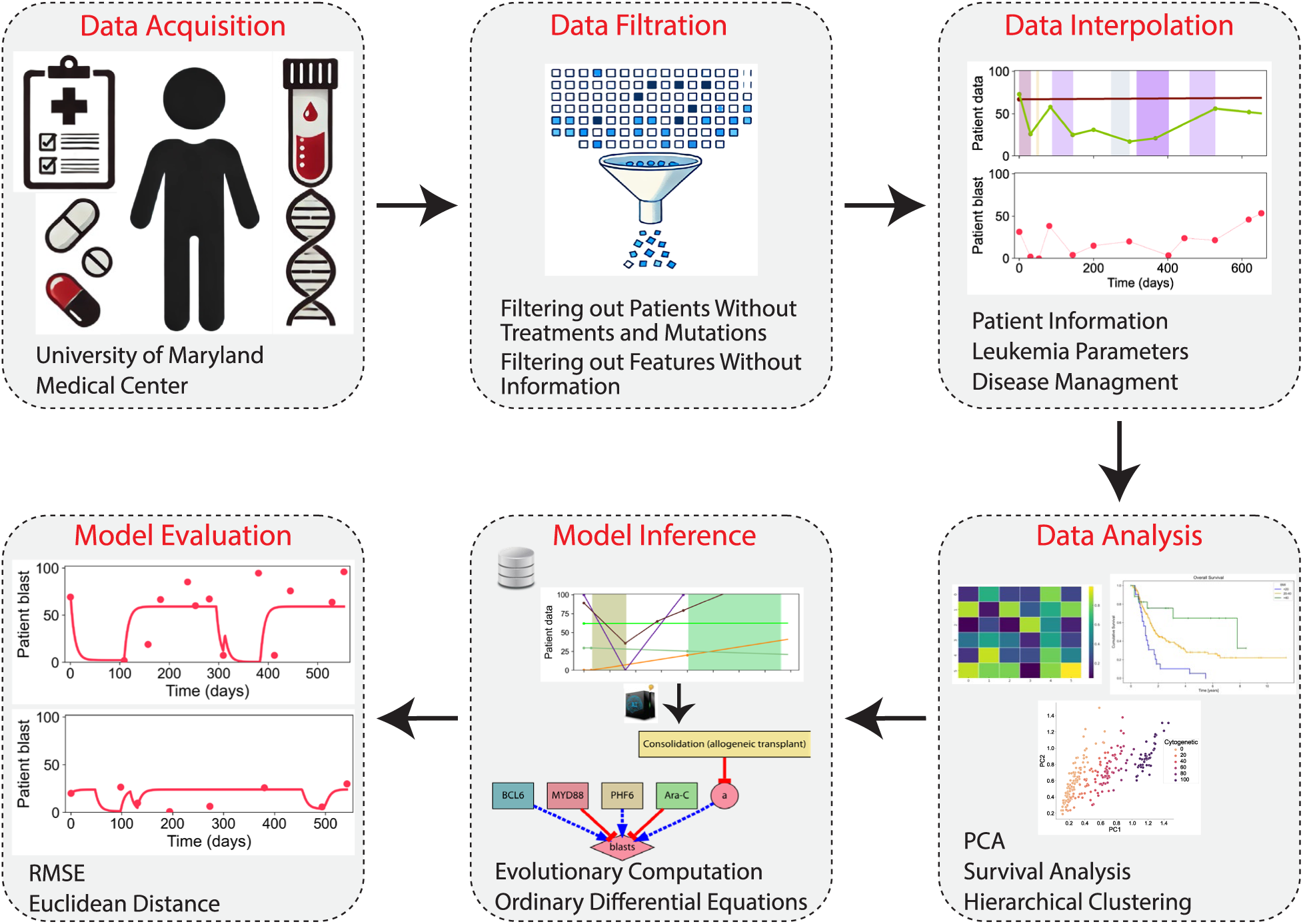
Pipeline for the inference of dynamic AML progression models using a novel interpretable machine learning framework. The pipeline consists of six stages: data acquisition (collecting clinical data, including patient information, genetic abnormalities, and treatment information), data filtration (removing patients and features with missing or insufficient information), data interpolation ( generating continuous temporal representations of multimodal clinical features), data analysis (applying techniques such as PCA, survival analysis, and clustering), model inference (discovering dynamic models with evolutionary computation and a system of differential equations), and model evaluation (assessing model performance with metrics such as RMSE and Euclidean distance).

### 2.1. Data collection and curation from an AML patient cohort

Data from patients with AML were collected from the University of Maryland Medical Center, comprising 467 patients and 125 multimodal variables, including patient, molecular, cytogenetic, and treatment data. Notably, each type of treatment was considered as a separate feature, but patients could receive combinations of treatments simultaneously or at different times throughout their clinical history. To ensure the integrity and quality of the analysis, a rigorous data-filtration process was implemented. 19 patients were excluded due to missing treatment information and 18 patients were excluded due to inconsistencies in the treatment dates. Additionally, 80 patients were excluded due to missing mutation information, and 145 patients were filtered out due to insufficient or missing data in a disease progression marker—blast percentages. Predictive features lacking informative values were then removed, resulting in the exclusion of 50 features from the dataset. After filtration, the final curated clinical dataset comprised a cohort of 245 patients with 75 predictive features, resulting in a robust and high-quality dataset for further analysis. A summary of the predictive features is presented in Table 1.

**Table 1.** Features in the curated multi-modal AML clinical dataset used for analysis and model training.

| Category | Number of Features | Features |  |
| --- | --- | --- | --- |
| Patient Information | 3 | Age, ECOG, BMI |  |
| Leukemia Parameters | 60 | Cytogenetic Abnormalities, Genetic Mutations (FLT3-ITD, FLT3-TKD, ASXL1, ASXL2, BCOR, BCORL1, BRAF, BRINP3, CALR, CBL, CEBPA, CSF3R, DDX41, DNMT1, DNMT3A, EED, ELANE, ETNK1, ETV6, EZH2, FLT3, GATA2, HNRNPK, HRAS, IDH1, IDH2, IKZF1, JAK2, JAK3, KIT, KMT2A, KRAS, MPL, NOTCH1, NPM1, NRAS, NSD1, PHF6, PRPF40B, PRPF8, PTPN11, RAD21, RUNX1, SETBP1, SF1, SF3A1, SF3B1, SH2B3, SMC1A, SMC3, SRSF2, STAG2, SUZ12, TET2, TP53, U2AF1, U2AF2, WT1, ZRSR2) |  |
| Disease management | 12 | Conventional Chemotherapy | Anthracycline-based, Ara-C, CLAG-M/FLAG-IDA, Cladribine-based |
|  |  | Drug Therapy | Hypomethylating agent (HMA), Asparaginase-based, FLT3 inhibitors (FLT3i), Venetoclax-based (VEN), IDH1/IDH2 inhibitors (IDH1/IDH2i) |
|  |  | Cellular therapies | Consolidation (allogeneic transplant), Donor lymphocyte infusion (DLI) |
|  |  | Others |  |

Figure 2 illustrates the dynamics of leukemia progression in two patients after integrating and interpolating patient information, leukemia-associated genetic parameters, and treatment interventions over time. The top panels for each patient depict longitudinal changes in clinical parameters (e.g., age, BMI, ECOG score), genetic mutations (e.g., FLT3-ITD, DNMT3A), and cytogenetic abnormalities, along with the timing and type of treatments, represented by shaded regions corresponding to specific therapeutic periods. The bottom panels show disease progression as the percentage of blasts, a key marker of leukemia. Patient A experienced an initial decline in blast percentage with Anthracycline and Ara-C (chemotherapy drugs commonly used to treat AML) treatments, followed by a relapse. Patient B progressed with more fluctuations in both cytogenetic abnormalities and genetic mutations, with periods of remission and progression aligned with treatment periods. These results highlight the complex interplay between individual patient factors, genetic and chromosomal abnormalities, and therapeutic responses in the prognosis and management of AML.

**Figure 2.**
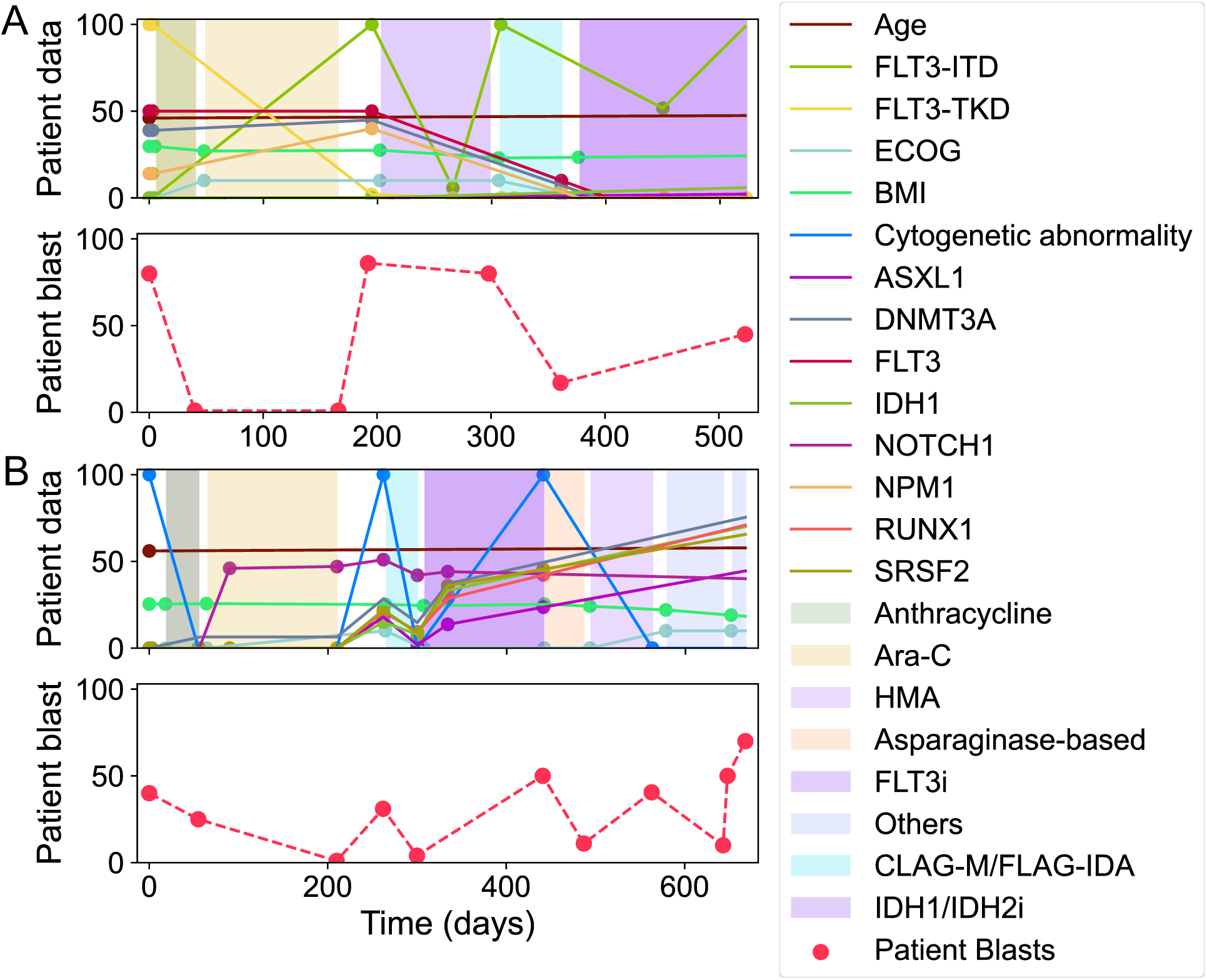
Longitudinal dynamics of cancer progression for two patients (A and B). Top panel (for each patient) illustrates longitudinal changes in clinical data, including patient characteristics, leukemia-associated genetic features, and treatment interventions. Shaded regions correspond to the treatment periods for each patient. Bottom panel (for each patient) represents the longitudinal dynamics of disease progression, measured as the percentage of blasts over time.

### 2.2. Analysis of the curated clinical dataset

We analyzed the key features curated in the clinical dataset to illustrate the overlap between critical mutations and treatment approaches in patients with AML. Figure 3A highlights the overlap in a selection of the key mutations that are most frequently observed across the patients. Each overlapping region indicates the number of patients with specific combinations of mutations across the entire clinical history of AML. Interestingly, six patients developed mutations in three genes concurrently: *DNMT3A*, *FLT3*-ITD, and *TET2*. Figure 3B shows the overlap among the four general classifications of the 14 treatment approaches included in the dataset. Each intersection in the Venn diagram shows the number of patients who received each combination of different treatment types during their clinical history of acute myeloid leukemia. Notably, 15 patients received all four treatment strategies.

**Figure 3.**
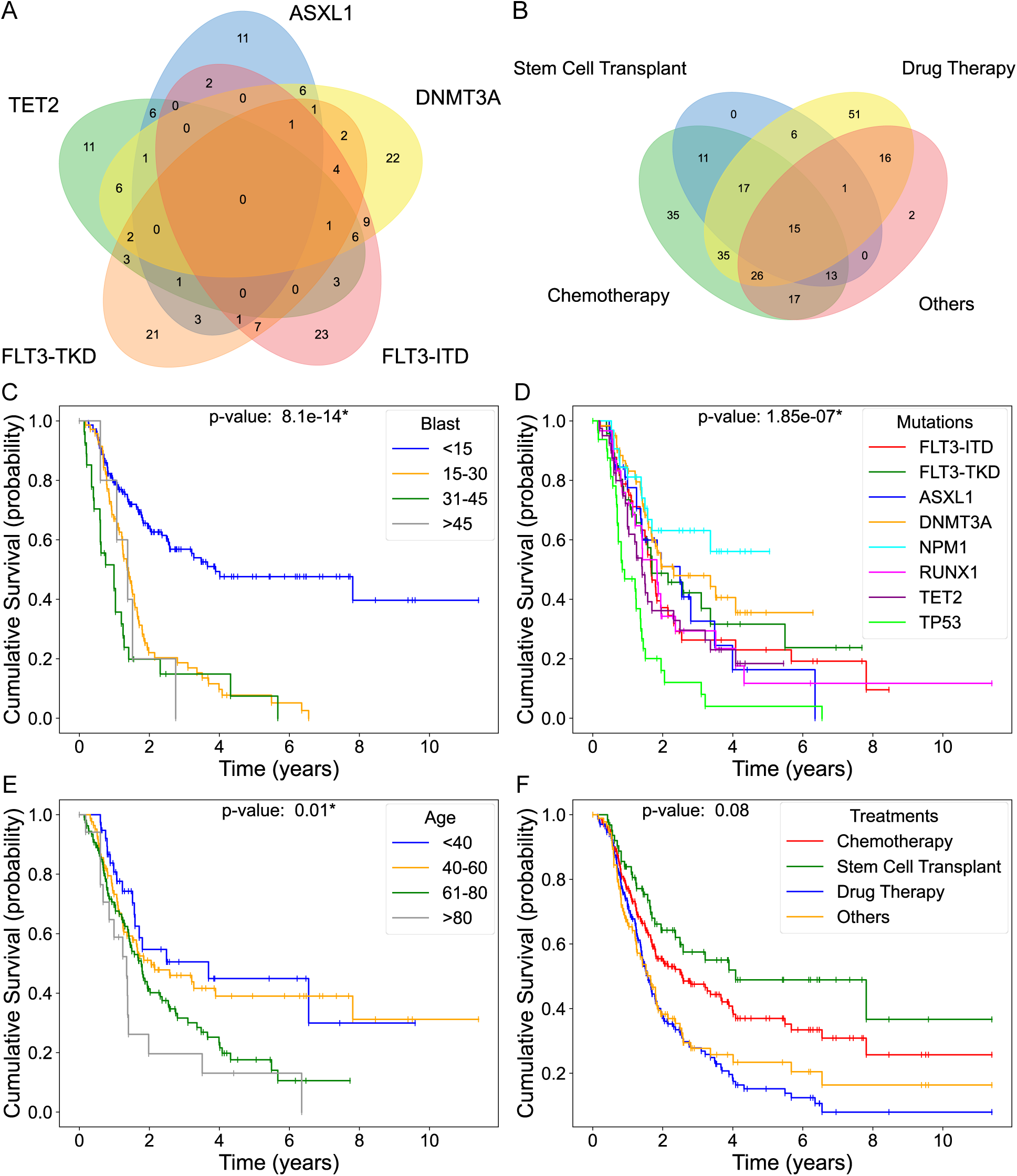
Venn diagrams and survival analysis of patient cohorts based on clinical, molecular, and disease progression features. **A.** Venn diagram illustrating the overlap of key genetic mutations in AML. **B.** Venn diagram showing the overlap among different treatment approaches. **C-F.** Kaplan-Meier survival curves illustrating cumulative survival stratified by age groups (C), blast percentages (D), gene mutations (E), and treatment regimens (F). The Venn diagrams as well as the treatment and mutation survival curves were generated using binary data. The age survival curves were stratified by patients’ age at the time of diagnosis. The blast curves were derived from continuous data, calculated as the area under the curve (AUC) divided by the observation duration (in days). Statistical significance among the survival groups was assessed using log-rank tests, with significance defined as *p* < 0.05 and indicated by *.

Next, we analyzed the curated dataset to reveal the cumulative survival curves of patients with different clinical features. Analysis of survival curves by blast counts revealed that patients with less than 15% blast counts had the highest survival probability; however, all groups with blast counts higher than 15% had significantly lower survivability, independent of their counts (Fig. 3C). Analysis of the survival probabilities by the main genetic mutations demonstrated a significant effect of different mutations on patient survival, where mutations in *TP53* (*p53*) resulted in the lowest survival probability (Fig. 3D). Figure 3E shows the survival curves by age, highlighting the significant decline in survivability of patients older than 61 years. Finally, figure 3F compares survival rates across different treatment types, showing that although their differences were not significant, stem cell transplantation resulted in the highest survival rates, followed by chemotherapy, drug therapy, and others.

To examine possible correlations between clinical features and disease progression (blast percentage), a principal component analysis (PCA) was performed on the curated AML clinical dataset. Figure 4 shows the PCA plot highlighting the selection of different features. From this analysis, we observed that patients with worse disease progression (high blast percentage) did not cluster separately from those with better prognosis (Fig. 4A). Instead, the main driver of the first principal component (PC1) was cytogenetic abnormality level (Fig. 4B); however, it did not correlate with the blast percentage (Fig. 4A). The main driver of the second principal component (PC2) was hypomethylating agents (HMA) (Fig. 4C), which did not correlate with the disease outcomes (Fig. 4A). The results showed that the remaining features did not cluster or correlate with disease outcomes (Fig. 4D-F). Additional analyses with non-linear dimensionality reduction (UMAP) resulted in similar non-clustered patterns for all features, except cytogenetic abnormalities and HMA, which did not correlate with blast percentages (Supplementary Figure 1). Similarly, a clustered heatmap including all features in the clinical dataset revealed no strong correlations between the different features or blast percentages (Supplementary Figure 2). Together, these results suggest that no strong clustering or correlation exists between clinical features and disease progression when analyzed using linear and nonlinear methods. This highlights the need for advanced computational approaches based on dynamic mathematical models to discover predictive mechanisms of AML disease progression from clinical data.

**Figure 4.**
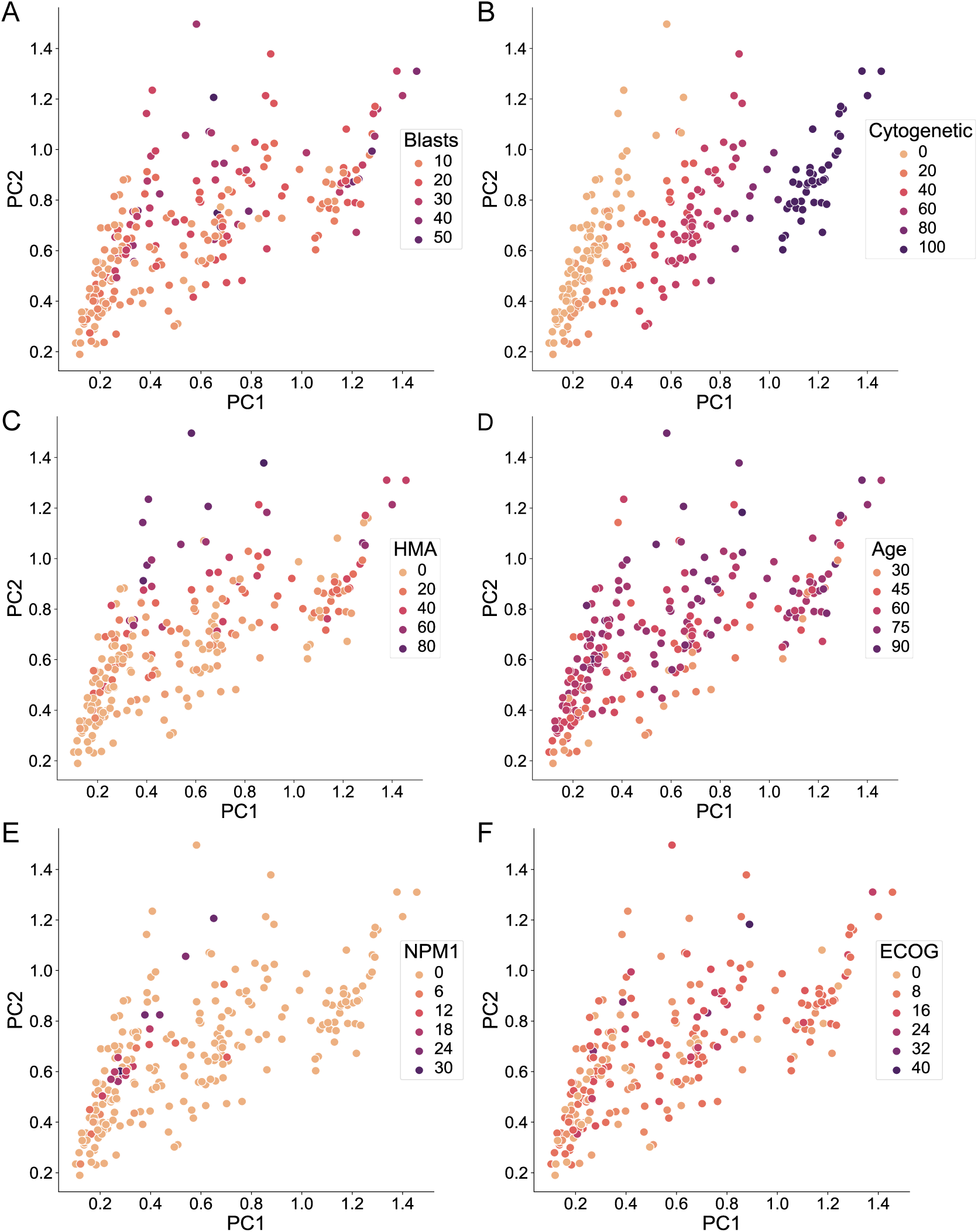
PCA plots exploring the relationships between clinical and molecular features and disease progression in AML. **A.** Blast percentage, representing a marker of disease progression. **B.** Cytogenetic abnormalities, reflecting changes in chromosome number or structure. **C.** Hypomethylating Agents (HMA), a class of drugs used in AML treatment. **D.** Age at time of diagnosis. **E.** Mutations in nucleophosmin 1 (*NPM1*), **F.** ECOG performance status, a functional measure of daily activity and self-care ability. For each feature, except age, data were derived from dynamic variables and calculated as the area under the curve (AUC) divided by the clinical history duration. Each point represents an individual patient, plotted according to their distribution across the first two principal components (PC1 and PC2). Points are colored by the respective clinical or disease feature to highlight potential correlations.

### 2.3. Machine learning for dynamic mechanistic models of disease progression

We developed an explainable machine learning method to automatically infer dynamic models of AML progression from patient clinical outcomes. The predictive models are formulated as a system of nonlinear ordinary differential equations (ODEs) to capture the temporal dynamics of disease progression. Each model includes three node types. Input nodes incorporate patient information, leukemia-associated genetic parameters, and treatments derived from the clinical data. Nodes in the middle layers process and integrate these input signals. The output node predicts how the disease progresses over time, as quantified by the percentage of abnormal cells (blasts). Links between nodes indicate nonlinear regulatory interactions from different features to the intermediate and output nodes. Such interactions include different numeric parameters specifying their regulatory strength and can be positive or negative and grouped in a necessary or sufficient fashion. Taking the interpolated clinical history of a patient as input, a model, when simulated, can precisely and quantitatively predict the temporal progression of the disease (blast percentage over the patient clinical history).

To infer an optimal interpretable model from the curated clinical dataset, the proposed machine learning approach leverages the power of evolutionary computation and high-performance computing. The algorithm starts with a random population of predictive models, each comprising input signals derived from the clinical data, an output node representing the predicted blast percentage, and a random number of intermediate nodes. In addition, random nonlinear regulatory interactions (excluding regulators for input signals) and random parameters are assigned to each initial model. These initial models are then translated into a system of ordinary differential equations and numerically simulated with the clinical data of each patient included in the training set to assess their ability to predict their disease progression dynamics as measured by an error function. Models that better recapitulate the observed disease progression (blast percentage over time) for the patients are preserved in the population, while those with worse errors are eliminated. The preserved models produce new offspring models through stochastic crossover (combining two parent models) and random mutations (changing parameters and adding or deleting links or nodes). These offspring models are subsequently simulated, scored, and integrated into the population for the next selection cycle. This iterative process continues until a model with zero error is identified, representing an accurate depiction of the temporal dynamics of disease progression. Notably, during this evolutionary process, the method automatically selects the most relevant features from the clinical dataset required to recapitulate disease progression, ensuring that only driver inputs are included in the final model.

### 2.4. Inference of dynamic mechanistic models of AML progression from clinical data

To infer mechanistic models of AML progression, the machine learning method was executed for 20 independent runs, each taking as input a random training subset of patients (90%) from the curated clinical dataset, with the rest (10%) used for testing. Figure 5A shows the overall evolutionary dynamics of the machine learning algorithm across the 20 independent runs. The results indicated that after approximately 24 h of computation, all the evolutionary processes converged, yielding models capable of recapitulating disease progression dynamics with zero fitness error. The performance of the models resulting from each run was evaluated using the root-mean-square error (RMSE). The models resulted in an average training RMSE of 9.48% and a testing RMSE of 11.62%. These results highlight the strong generalization capability of the method, as evidenced by the minimal difference between the training and testing RMSE values. The average model complexity (number of nodes and links) was 57, achieving a balance between capturing the dynamic dataset and maintaining interpretability.

**Figure 5.**
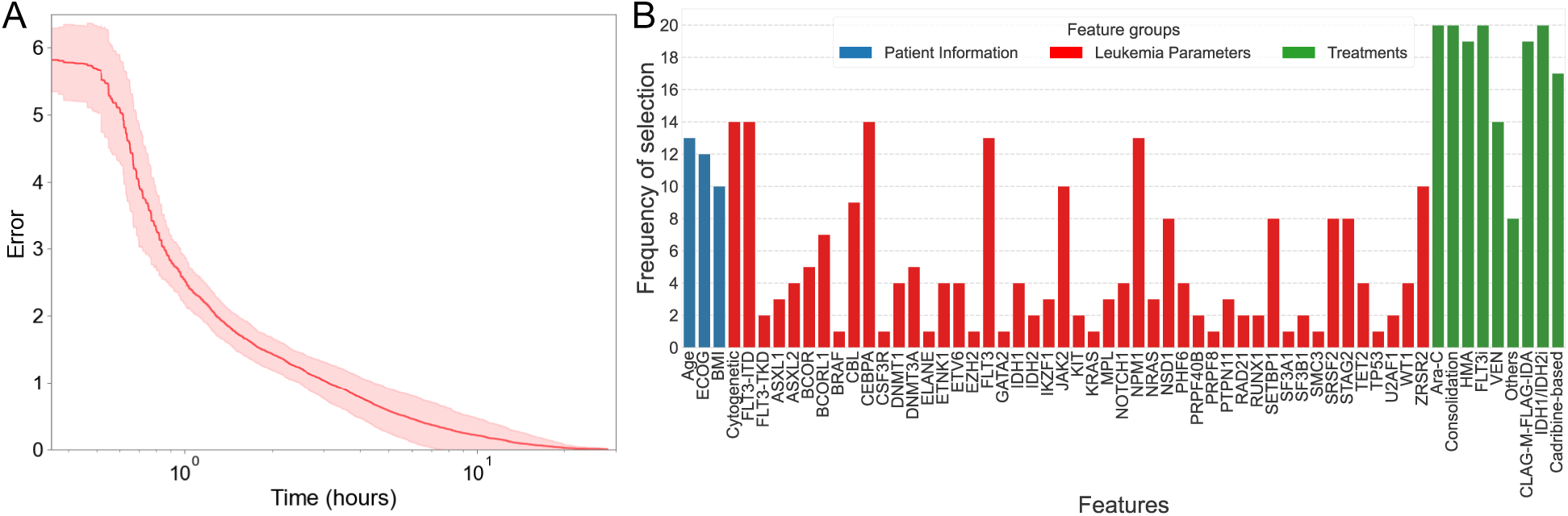
Overview of evolutionary dynamics and feature selection frequency over 20 independent runs of the machine learning methodology. **A.** Error trajectory of the best-performing model on the training dataset. The fitness error decreases consistently, converging to zero after approximately 24 hours of execution. The line and shaded area indicate the average and standard deviation, respectively, of the error values across all runs. **B.** Frequency of feature selection across all runs. For each run, the training data was generated by randomly shuffling the dataset and selecting 90% of the samples.

Figure 5B illustrates the frequency of feature selection across the 20 runs of the proposed methodology, identifying the predictive features that significantly contribute to the dynamics of AML disease progression. Notably, chemotherapy (Ara-C, other than high dose-Ara-C), consolidation therapy (high dose Ara-C), FLT3 inhibitors (FLT3i), and IDH inhibitors (IDH1/IDH2) were selected to be included in the inferred model for every run, emphasizing their critical importance for AML management. Additionally, the predicted features included other key factors, such as age, ECOG performance status, and cytogenetic abnormalities, as well as *FLT3*- ITD, *CEBPA*, and *NPM1* mutations, as primary drivers of AML progression. Overall, this analysis revealed the most relevant features influencing AML progression, as identified by the inferred predictive models.

### 2.5. Explainable machine learning insights into AML progression and patient-specific dynamics

Analysis of the inferred regulatory effect of the selected features across all runs on blast percentages revealed the predicted drivers of AML disease progression. Figure 6A illustrates the mechanistic regulation strength of each feature over the 20 independent runs of the predictive methodology. Each bar represents a feature’s overall impact on the blast percentages computed as the normalized link strength (value computed as the average of the half-response parameters and sign as the multiplicative positive or negative effect in a pathway) across all predictive models. The results showed that nearly all treatments together with ECOG performance were identified by the predictive models as having a strong negative effect on blast percentages, indicating a positive correlation with improved AML patient outcomes. Conversely, cytogenetic abnormalities, as well as mutations in *FLT3*-ITD, *CEBPA*, *NPM1*, *ZRSR2*, and *FLT3*, were predicted to have a strong positive effect on blast percentages, suggesting their roles as major drivers of AML progression. Overall, this analysis highlighted the predicted effects of link strength from predictive features on the blast percentages, as identified by the models.

**Figure 6.**
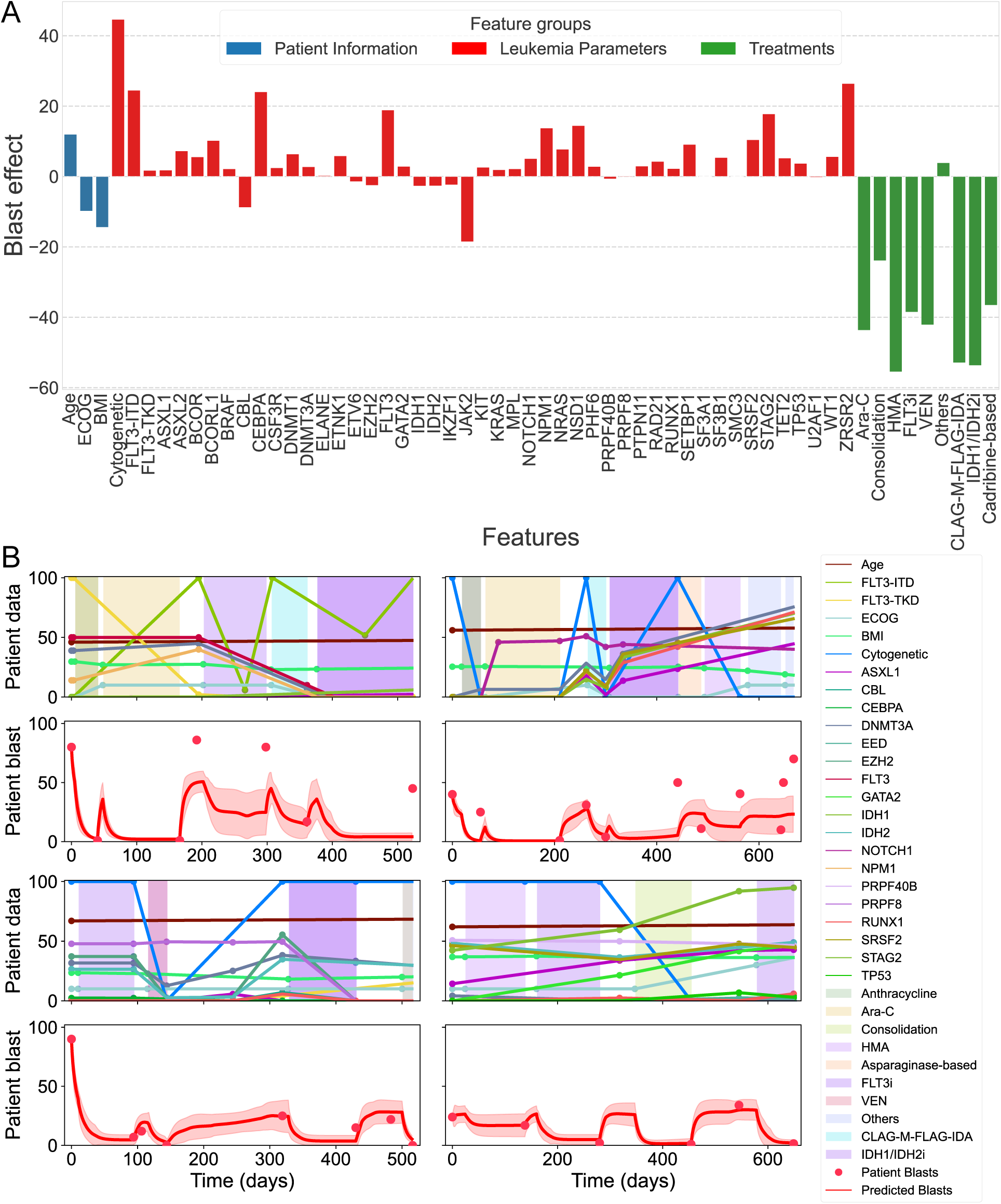
Overview of clinical feature effects on blast percentages and predicted patient blast dynamics from the inferred mathematical models. **A.** Predicted mechanistic regulation effects of clinical features on blast percentages. **B.** Predicted disease progression dynamics for four patients in the curated dataset. The line and shaded region represent the average and standard deviation, respectively, of error values across all models.

Figure 6B illustrates the overall disease dynamics of the models, averaged across the 20 independent runs, for four individual patients from the curated dataset (see Supplementary Figure 3 for all patients). The patient-specific blast dynamics highlight both the consistency and variability of predictions across different runs over time. The observed trends demonstrate how all models effectively captured the dynamics of individual patient blast percentages while adapting to the variability introduced by training data randomization and stochastic model initialization. These findings provide valuable insights into disease progression, laying the groundwork for the development of tailored patient-specific predictive strategies.

Figure 7 illustrates the mechanistic regulation in the best-performing dynamic model inferred using the proposed predictive methodology, together with its simulation with two different patients (see Supplementary Information for the model system of equations). As shown in Figure 7A, the discovered model has a complexity of 47 and includes three intermediate nodes to integrate the different mutation and treatment effects into the blast dynamics. Figure 7B-C demonstrates the model simulations for two patients from the training and test datasets, respectively, highlighting its accuracy in capturing both observed and unseen patient dynamics. These results indicate that the proposed predictive methodology can not only accurately model the progression of AML but also generalize well to new patient data, demonstrating its ability to understand and forecast disease dynamics in clinical applications.

**Figure 7.**
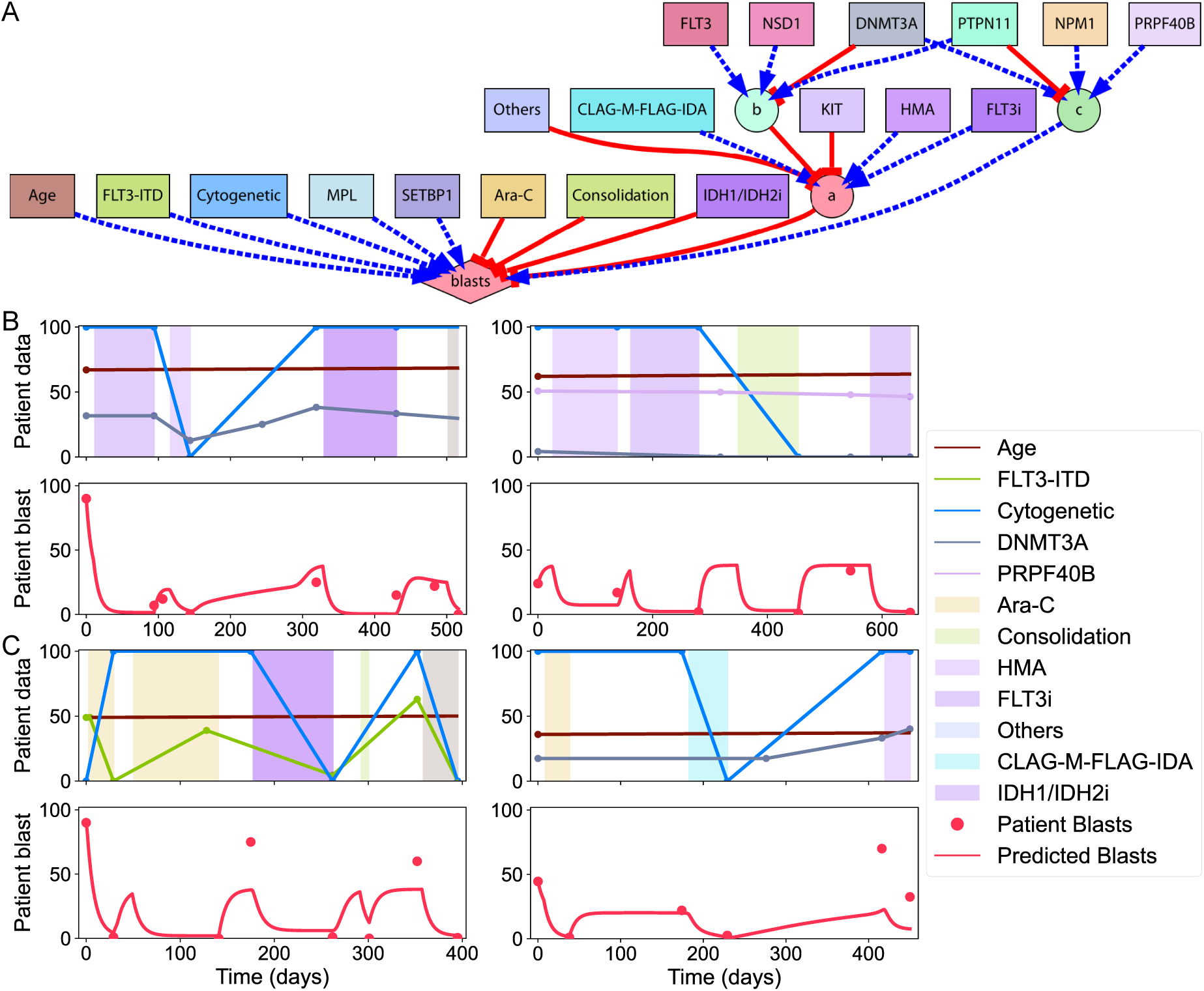
Inferred model dynamics for AML disease progression. **A.** The dynamic model inferred by the predictive methodology incorporates input features (squares), intermediate variables (circles), and an output (diamond) node. Regulatory interactions are represented as either positive (pointed arrows) or negative (blunt arrows) and grouped as necessary (solid lines) or sufficient (dashed lines). **B-C.** Simulations of the best model. (B) shows two patients from the training dataset, highlighting the predicted blast percentages over time, while (C) depicts two patients from the test dataset, highlighting model performance on unseen data. Curve, line, and shaded area colors in (B) and (C) correspond to the node colors in the model network diagram in (A).

## 3. Discussion

In this study, we collected the clinical histories of 467 patients diagnosed with AML at the University of Maryland Medical Center, including 125 multimodal features recorded between the years 2006-2021. Following rigorous filtration and data interpolation, the curated, unique dynamic dataset comprised 245 patients with 75 clinical predictive features, including patient demographics, genetic and chromosomal abnormalities, disease management strategies, and progression markers (blast percentages), for downstream analyses. Survival curves of different patient cohorts revealed that survival rates were lower among older patients, highest among those with less than 15% blasts, and most favorable for patients receiving stem cell transplant treatments. However, correlation and clustering analyses identified no strong relationships between clinical features and disease progression, as evaluated using both linear and nonlinear methods. These findings highlight the complex interplay between clinical features and AML progression and underscore the need for advanced computational approaches to better understand these complex relationships.

Hence, we sought to identify the mechanisms underlying AML progression by developing a novel predictive methodology to discover dynamic mathematical models from longitudinal clinical data. A high-performance explainable machine learning approach, together with a dynamic mathematical modeling framework, was then developed and applied to the curated clinical dataset, enabling efficient inference of models capable of predicting disease progression. The resulting models, represented as systems of ordinary differential equations (ODEs), included essential gene mutations, regulatory links, and all numeric parameters, allowing for simulations of the temporal dynamics of AML progression. The discovered models with the machine learning methodology showed minor variations in predicting disease progression while maintaining strong generalization power to accurately capture disease dynamics in novel patients. Additionally, the discovered models identified key predictive markers and elucidated their mechanistic effects on disease progression, highlighting drivers or mitigators of AML. These insights hold significant potential for improving the diagnosis and treatment of AML in clinical and research settings.

The proposed methodology was applied to a comprehensive cohort of AML patients, yet it has the potential to be extended to predict treatment outcomes for other acute diseases. Regarding its efficiency, the method begins with random initial models, but as a possible optimization, it could incorporate an initial population of models including pre-established nodes and interactions from known knowledge relevant to disease progression. Incorporating different interpolation techniques could further improve the methodology by better capturing the dynamics of disease progression, allowing for a more accurate reconstruction of temporal dynamics in the data. Additionally, the algorithm is currently designed to return a single model per run, although multiple models could provide a more accurate representation of disease progression dynamics. Future work will focus on extending the methodology with evolutionary multi-objective and diversity-preserving algorithms (45) to enable the discovery of a broader set of models—an atlas of regulatory models—capable of predicting AML disease progression.

## Acknowledgements

We thank the members of the Lobo Lab for helpful discussions. This work was supported by the National Institute of General Medical Sciences of the National Institutes of Health under award number R35GM137953. The content is solely the responsibility of the authors and does not necessarily represent the official views of the National Institutes of Health. Computations used the UMBC High Performance Computing Facility (HPCF) supported by the NSF MRI program grants CNS-1920079 and OAC-1726023.

## Data Availability

The source code for the machine learning, simulation, and visualization methods are freely available on GitHub (https://github.com/lobolab/kinetic-leukemia).

## Supplementary Information

### 1. Materials and Methods

We developed a computational pipeline for the analysis and mechanistic inference of AML progression dynamics from multimodal clinical data, including patient information, leukemia parameters, and disease management strategies. The proposed pipeline comprises several key steps: data curation, data filtration, data interpolation, data analysis, model inference, and model evaluation (Figure 1).

#### 1.1. Data acquisition

For this study, an anonymized clinical dataset was curated from the University of Maryland Medical Center comprising 467 patients diagnosed with acute myeloid leukemia between October 2006 and June 2021 (protocol approved by the University of Maryland, Baltimore Institutional Review Board and the University of Maryland, Baltimore County Office of Research Protections and Compliance). The patients’ ages ranged from 18 to 98 years and comprised a male/female ratio of 1.33. This comprehensive clinical dataset contains detailed, temporal patient information, leukemia-associated genetic parameters, molecular and cytogenetic data, disease management, including treatments applied to each patient, and a disease progression marker (blast percentages). Genetic mutations were measured using either next-generation sequencing (NGS) or PCR-based fragment analysis. Blast percentages were determined based on bone marrow aspirate and biopsy and if not done, then from peripheral blood. Patient information was collected from the date of diagnosis to the date of death or last follow-up. All therapies administered and the response to therapy using ELN 2017 response criteria were collected (46). Moreover, blast percentage, cytogenetic exam and molecular testing using NGS were collected at each response examination, when available. Data was stored in REDCap and was updated till 3/2022 (47).

#### 1.2. Data filtration

The data curation process involved a rigorous filtration approach to ensure consistency, accuracy, and quality of the collected data for downstream analyses. Patients lacking data on treatments, with inconsistencies in treatment dates, missing genetic mutation data, or fewer than two data points in the disease progression marker (patient blasts) were excluded. In addition, clinical features with missing information were excluded to improve model interpretability and reduce computational complexity. This thorough curation of the clinical dataset ensured robust and accurate analyses, while improving the efficiency and performance of the machine learning methodology to infer predictive models.

#### 1.3. Data interpolation

Discrete patient, mutation, and disease progression data points were transformed into dynamic continuous representations using a linear interpolation approach. In this way, linear curves were computed between discrete data points to generate a continuous temporal representation of patient trajectories, while preserving the underlying trends in clinical data. Genetic mutations in the collected dataset were recorded as the percentage of allele frequency (AF), defined as the proportion of sequencing reads containing a specific mutation relative to the total reads at the corresponding genomic location. These percentages provide a quantitative measure of mutation burden within each sample. In the collected data, two types of blast measurements were available: peripheral blood (PB) blasts and bone marrow (BM) blasts. Due to the limited availability of these values across time, we applied a strategy to combine them. Specifically, when both PB and BM blast values were clinically recorded at the same time point, their average was calculated and used as the representative blast value. Averaging these values helped create a smoother and more consistent transition between time points, reducing abrupt changes and improving the continuity of blast percentage trajectories for modeling. If only one type was available at a given time, that value was directly used as the disease representative blast percentage for that time point. This approach facilitated the generation of a more continuous and comprehensive blast trajectory for each patient, effectively minimizing the impact of missing data and enhancing the accuracy of longitudinal analyses to better understand AML progression. Cytogenetic abnormality data were categorized and transformed into binary continuous temporal variables. Labels indicating significant cytogenetic abnormalities (e.g., abnormal metaphase findings, FISH abnormalities, persistent abnormalities, and partial remission) were assigned a numeric value between 0 and 100. Treatment intervention data points were transformed into binary (a particular treatment being applied or not) continuous temporal functions. Such temporal binary data provided a clear differentiation between the treated and untreated periods for each type of disease management intervention, allowing the model to accurately assess the impact of each clinical intervention on AML disease progression. By integrating and normalizing these diverse temporal data points, we constructed a longitudinal continuous multimodal dataset that could serve as an input for inferring mechanistic dynamic models capable of predicting AML disease progression.

#### 1.4. Data analysis

After filtering, the curated multimodal AML dataset was analyzed to assess their relationships and predictive ability with respect to the disease progression marker, blast percentage. Venn diagrams were constructed by transforming mutation and treatment strategies into binary data (occurrence or absence during the clinical history of a patient) and visualizing their overlap among all patients. Cumulative survival plots were generated using the Kaplan-Meier method (48), grouping patients by age, genetic mutations, treatment interventions, and disease progression markers. For the age feature, survival analysis was based on the age at diagnosis, while mutation and treatment strategies were assessed as binary data (occurrence or absence during the clinical history of a patient). The disease progression marker (blast percentages) was analyzed using continuous values calculated as the area under the curve (AUC) normalized by the clinical history duration for the patient. Statistical significance between survival groups was computed using a log-rank test.

To investigate the correlations between patient features and disease progression markers, dimensionality reduction and clustering techniques were applied. Principal Component Analysis (PCA) and Uniform Manifold Approximation and Projection (UMAP) were applied to identify correlations between the clinical features and the diagnostic marker. Additionally, a heat map was generated to provide a detailed overview of feature clustering and its association with disease progression. PCA, UMAP, and heatmap analyses were performed using continuous values derived by calculating AUC of features normalized by the clinical history duration.

#### 1.5. Model inference

An explainable Machine Learning (ML) method was developed using high-performance evolutionary computation to efficiently discover dynamic models, including the number of nodes, their regulatory interactions, and parameters derived from the clinical dataset. The method can infer the topology and parameters of a dynamic predictive model defined by a system of ordinary differential equations, which can accurately simulate disease progression, defined as the blast percentage over time. Taking the longitudinal multimodal dataset curated from patients as input, the method can capture the temporal dynamics of AML progression, providing a reliable and interpretable framework for understanding and predicting disease trajectories.

The ML approach leverages evolutionary computation to automatically derive a dynamic model that recapitulates the disease progression dynamics. The algorithm iteratively evolves a population of candidate predictive models through reproduction, fitness assessment, and selection, ultimately returning an optimal model that best represents the underlying dynamics. An island distribution strategy was employed to enhance parallel processing, while promoting robustness and diversity among candidate models. New models are generated through stochastic combinations of existing models in addition to random mutations in the parameters, interactions, and nodes in the model. A crossover operator creates two child models by randomly merging models from two parents within the population, redistributing nodes, and regulatory interactions without duplicating nodes or altering the kinetic parameters. Subsequently, a mutation operator is applied, which may add or remove nodes and regulatory interactions as well as modify parameters within predefined ranges sampled from a uniform random distribution. The output node (blast percentage) is always preserved. The algorithm is biased towards simpler models by assigning a higher probability of deletion mutations compared to duplication mutations, thereby preventing model bloating (49).

The selection mechanism is based on deterministic crowding (50), replacing new offspring only if they have equal or improved fitness compared with their closest parent models. The algorithm iterates until it finds a model with zero error, after which it continues until 2000 additional generations are completed without any reduction in the model complexity (number of edges plus number of nodes). The machine learning method was run with the following meta-parameters: crossover rate, 75%; mutation rate, 1%; link/gene duplication rate, 1%; link/gene deletion rate, 1.5%. Each run used 32 subpopulations (islands), with each consisting of 64 individuals. To enhance diversity and exploration in the search space, islands were randomly paired, and their models were randomly swapped every 250 generations.

A disease progression model simulator was developed to predict patient longitudinal dynamics using a system of ordinary differential equations (ODEs). Models comprise nodes representing the dynamic levels of the patient, mutation, treatment, and disease progression data, together with intermediate nodes integrating these complex signals. Hence, nodes in a model are categorized into three groups: (1) input nodes representing patient information, genetic mutations and abnormalities, and treatment interventions that follow the dynamic continuous values in a patient as curated in the clinical dataset; (2) intermediate nodes representing integration steps of signal dynamics; and (3) the disease progression output node predicted by the model as the blast percentage over time. A model also includes all the regulatory interactions between the nodes and defines the type and parameters of each interaction. Input nodes can regulate other nodes but cannot be regulated by either themselves or other nodes, as they follow the dynamics interpolated from the patient’s clinical data. In contrast, intermediate nodes and the output node can be regulated by other nodes and themselves, as they change their values according to the equations defined in the model and evolve with the machine learning methodology.

Each intermediate node and output node in the predictive model includes two parameters: production and decay rates. Regulatory interactions are characterized as either positive or negative and modeled using a Hill function with two parameters: the Hill coefficient and half-response values. Nodes can receive regulations from multiple nodes simultaneously, and these regulatory inputs can be classified as either necessary or sufficient. Necessary positive regulations are combined using a multiplication operator, whereas sufficient positive regulations use both multiplication and summation. Negative regulations are combined exclusively using multiplication operators. This approach provides a robust and interpretable mechanism for predicting AML progression dynamics by incorporating patient-specific factors and dynamic regulatory interactions.

Thus, the level *n*_*i*_ of an intermediate or output node *i* is given by

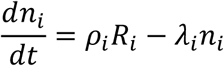

where *ρ*_*i*_ is the production constant and *λ*_*i*_ is the decay constant. The regulatory term *λ*_*i*_ is modeled using a Hill function for each regulatory interaction that affects the node *n*_*i*_. For example, the following equation describes a regulatory term *λ* affecting the node *n*_*i*_ when it is regulated by two sufficient positive nodes *r* and *g*, two necessary positive nodes *p* and *y*, and a negative node *o*:

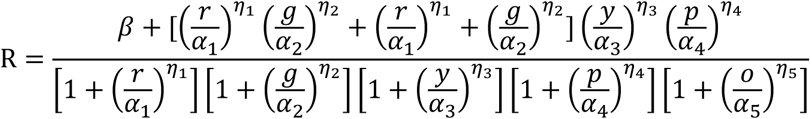

where *η*_*i*_ are the Hill coefficients, and *α*_*i*_ are the half-response values. Model parameter ranges were as follows: Hill coefficient (1,10), half-response value (0.01,100), decay constant (0.1, 1), and production constant (0, 100).

#### 1.6. Model evaluation

The fitness of a model measures its capability to accurately predict disease progression dynamics (blast percentage) when simulated using the clinical data of each patient in the training set, as quantified through an error function. The error for a candidate model is calculated by simulating the disease dynamics for each patient in the dataset and computing the average Euclidean distances between the patient data and predicted blast percentage values across the entire clinical history of each patient. To minimize overfitting and increase the discovery of simpler models, two parameters, represented by the local threshold *α* and global threshold *β*, are included in the error function. These thresholds ensure that models with concentration scores below these limits are assigned an error of zero. This strategy avoids adding unnecessary complexity to models, so that extra intermediate nodes or regulatory interactions are included only if they significantly improve predictive performance. In this way, the error of a model with respect an input dataset comprising the curated clinical data of a set of patients is calculated as

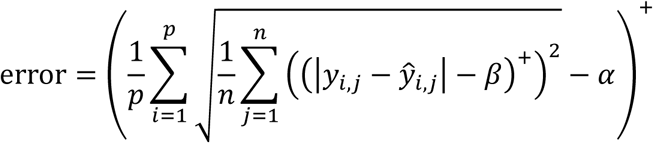

where *p* and *n* are the number of patients and the number of data points for the blast marker in patient *i*, respectively, *y*_*i*,*j*_ and *ŷ*_*i*,*j*_ are the recorded patient and model-predicted blast values at time point *j* for patient *i*, *α* and *β* are the local and global thresholds, respectively. For all the runs, the *α* and *β* threshold parameters were set to 10 and 1, respectively. The function (*x*)^+^represents the positive part function, which outputs 0 if *x* is negative and *x* if *x* is nonnegative.

In addition, the root-mean-square error (RMSE) was used to evaluate the model performance on the testing set by comparing the observed patient blast values with the model’s predicted values. A lower RMSE value indicates a closer fit between the predicted and actual blast values, reflecting a higher accuracy of the model’s predictions. The RMSE is calculated as

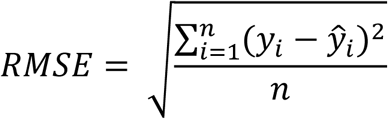

where *n* is the number of data points, and *y*_*i*_ and *ŷ*_*i*_ denote the observed and model-predicted values at the time point *i*, respectively.

#### 1.7. Implementation

Data analysis and visualization were conducted using a diverse set of Python libraries. Survival analysis was conducted using the *lifelines* library, while hierarchical clustering was carried out using the *scipy* library. The *venn* library was used to generate the Venn diagrams. Dimensionality reduction techniques, including Principal Component Analysis (PCA) and Uniform Manifold Approximation and Projection (UMAP), were implemented with the *scikit-learn* library. All visualizations were created with the *seaborn* and *matplotlib* libraries, providing clear and effective graphical representations of the results. We implemented the machine learning and model simulation methodology in C + + with the standard Eigen (Gaël Guennebaud, Benoît Jacob, and others), Qt (The Qt Company Ltd.), and Qwt (Uwe Rathmann and Josef Wilgen) libraries. A generalized Runge-Kutta eighth-order solver with an adaptive step size (51) was developed to numerically solve the system of ordinary differential equations (ODEs). The implementation leveraged 64 parallel threads and was executed on a server equipped with two 32-core Intel Xeon Gold 6548Y+ CPUs and 512GB of RAM to assess its performance.

### 2. Supplementary Figures

**Supplementary Figure 1.**
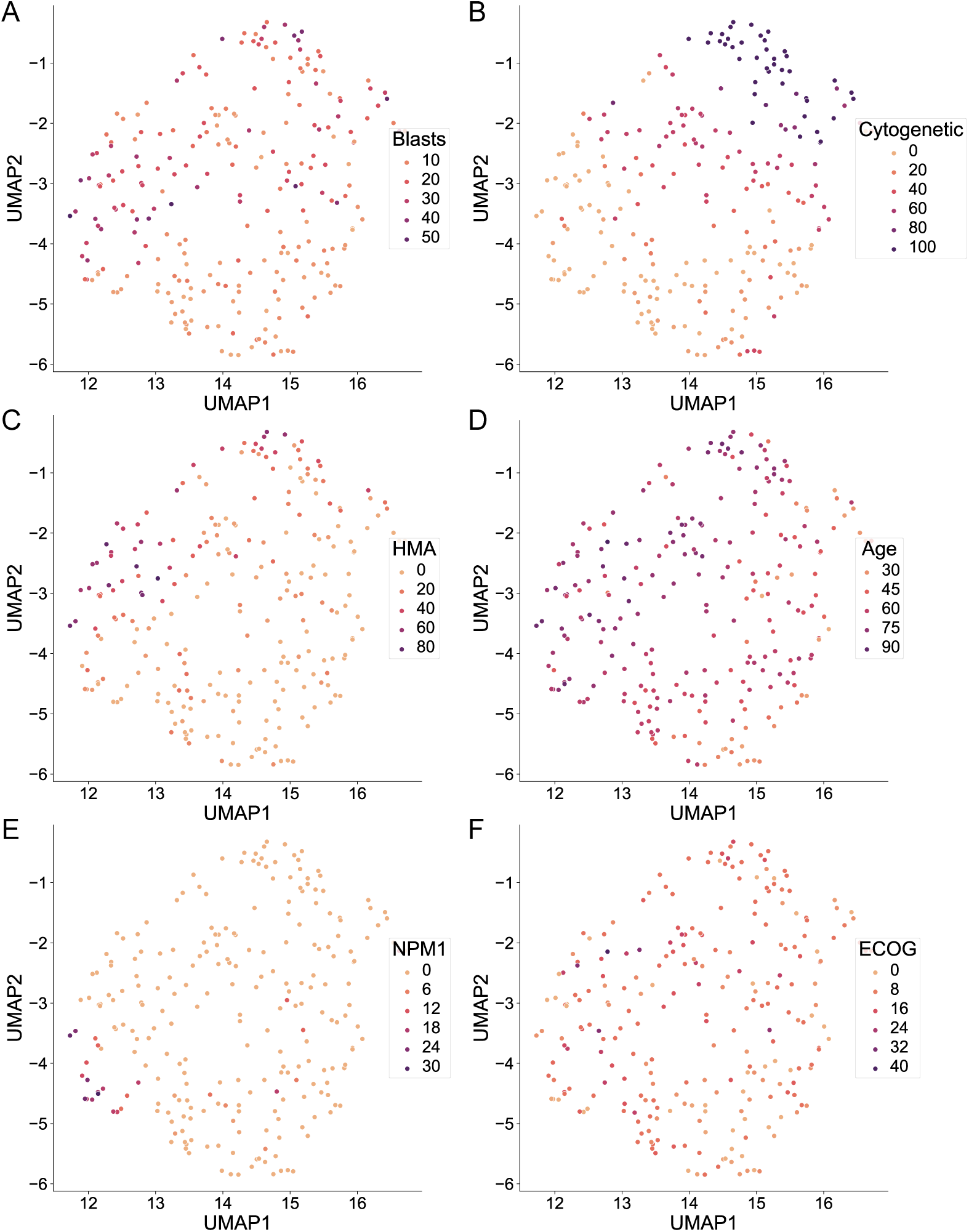
UMAP plots highlight the relationships between clinical, molecular, and disease progression features in the curated AML patient dataset. **A.** Blast percentage, representing the marker of disease progression. **B.** Cytogenetic abnormalities, reflecting changes in chromosome number or structure. **C.** Hypomethylating Agents (HMA), a class of drugs used in AML treatment. **D.** Age at time of diagnosis. **E.** Mutations in *NPM1*, **F.** ECOG performance status, a functional measure of daily activity and self-care ability. For each feature, except age, data were derived from dynamic variables and calculated as the area under the curve (AUC) divided by the clinical history duration. Each point represents an individual patient, plotted according to their distribution across the first two UMAP dimensions. Points are colored by the respective clinical or disease feature to highlight potential correlations.

**Supplementary Figure 2.**
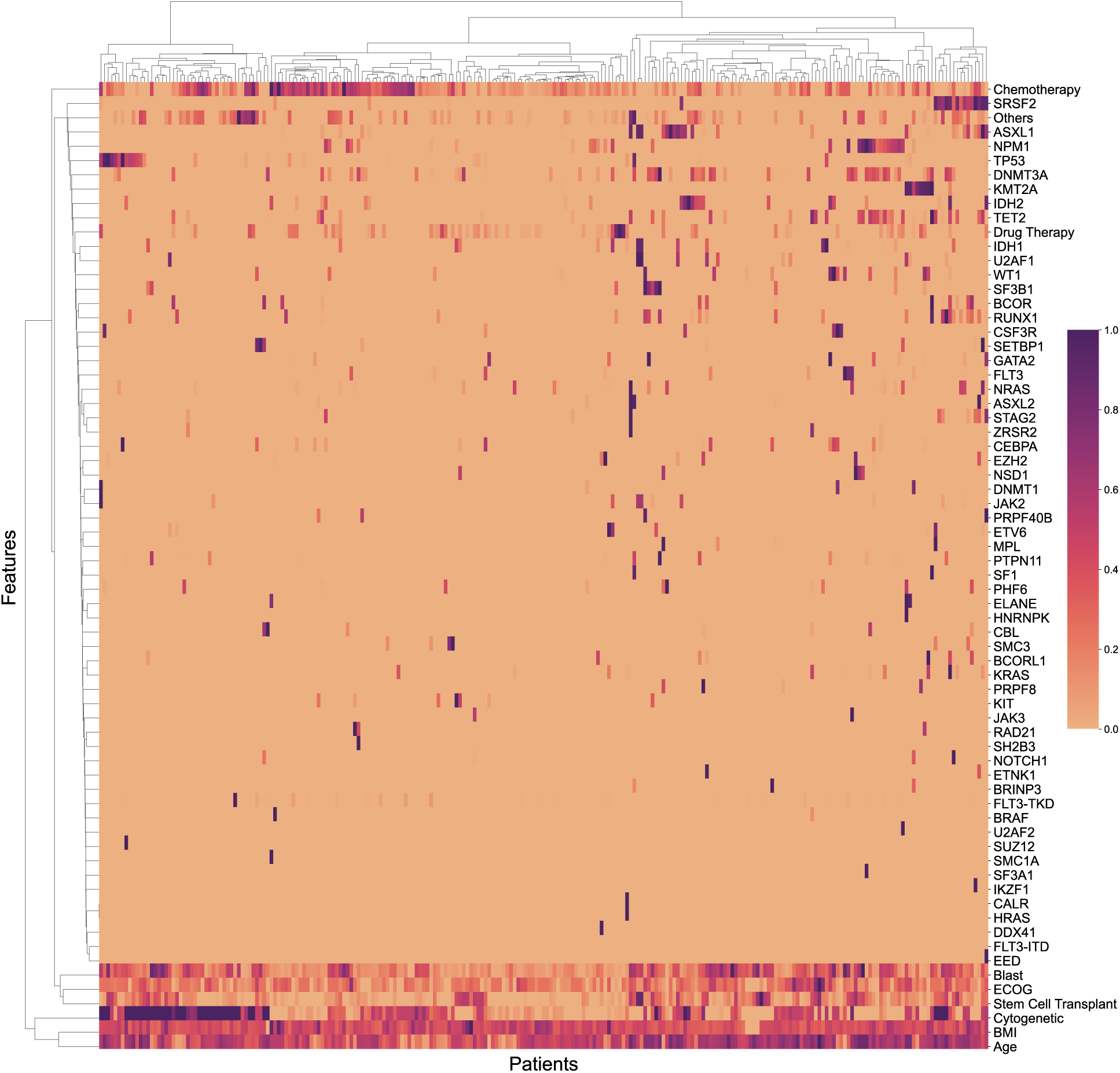
Clustered heatmap of patients based on all clinical, molecular, and disease progression features. For each feature, data were derived from continuous variables, calculated as the area under the curve (AUC) divided by the clinical history duration. The color indicates normalized values across the cohort for each feature, with dark and light colors representing high and low values, respectively.

**Supplementary Figure 3.**
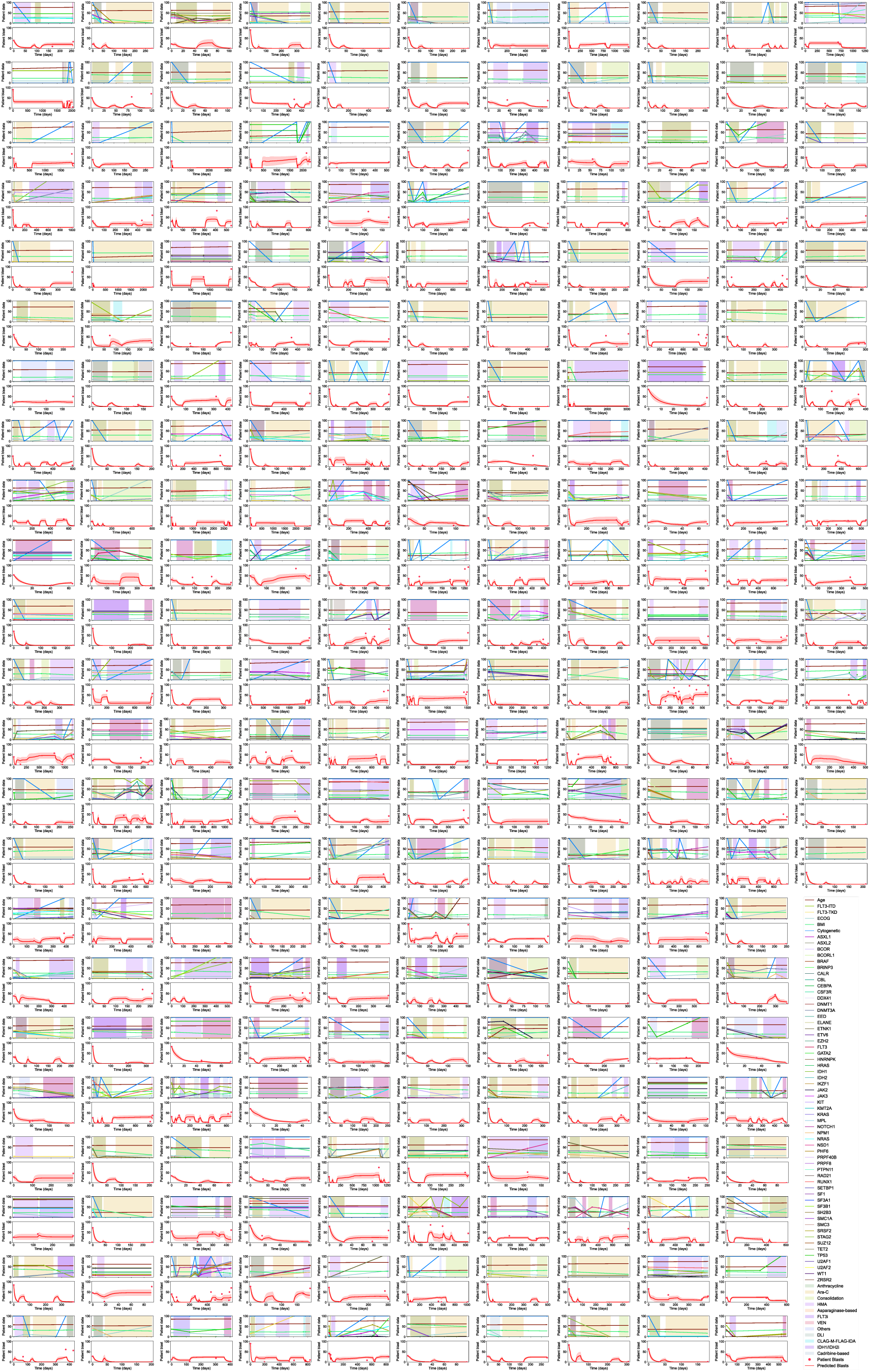
Predicted disease progression dynamics for all patients in the curated dataset. The line and shaded region represent the average and standard deviation, respectively, of error values across all models.

### 3. System of equations governing the model illustrated in Figure 7a

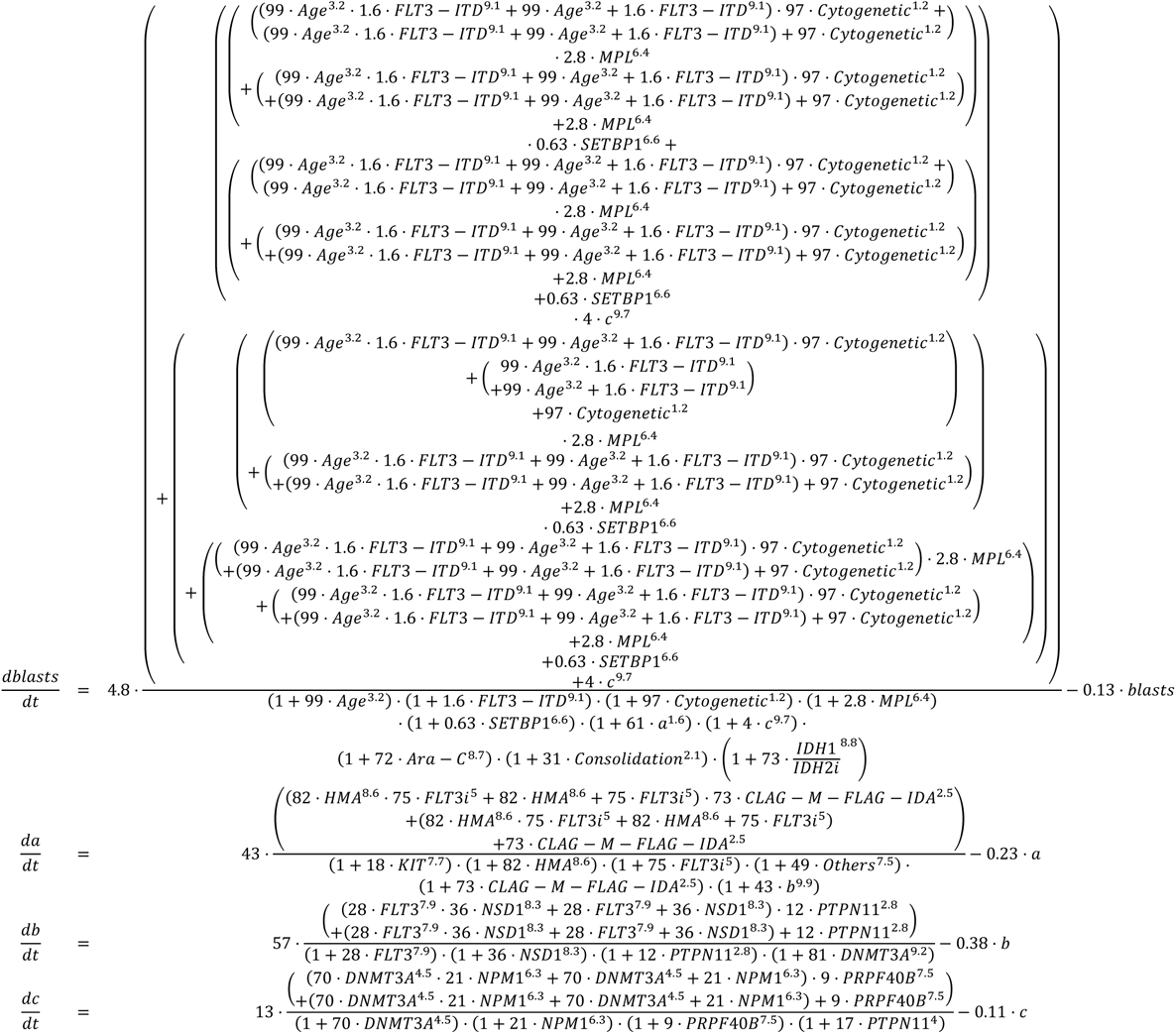

